# Coffee intake is associated with telomere length in severe mental disorders

**DOI:** 10.1101/2025.09.25.25336622

**Authors:** Vid Mlakar, Marta Di Forti, Els F. Halff, Deepak P. Srivastava, Ibrahim Akkouh, Srdjan Djurovic, Carmen Martin-Ruiz, Daniel S. Quintana, Viktoria Birkenæs, Nils Eiel Steen, Monica B. E. G. Ormerod, Ole A. Andreassen, Monica Aas

**Affiliations:** Social, Genetic and Developmental Psychiatry Centre, Institute of Psychiatry, Psychology and Neuroscience, King’s College London, London, UK; Department of Psychosis Studies, Institute of Psychiatry, Psychology and Neuroscience, King’s College London, London, UK; Department of Basic and Clinical Neuroscience, Institute of Psychiatry, Psychology and Neuroscience, King’s College London, London, UK; MRC Centre for Neurodevelopmental Disorders, King’s College London, London, UK; South London and Maudsley NHS Foundation Mental Health Trust, London, UK; Department of Medical Genetics, Oslo University Hospital, Oslo, Norway; Department of Psychology, University of Oslo, Oslo, Norway; NevSom, Department of Rare Disorders, Oslo University Hospital, Oslo, Norway; Centre for Precision Psychiatry, Division of Mental Health and Addiction, University of Oslo, Oslo, Norway; Section for Clinical Psychosis Research, Division of Mental Health and Addiction, Oslo University Hospital, Oslo, Norway; Department of Psychiatric Research, Diakonhjemmet Hospital, Oslo, Norway; BioScreening Core Facility-CAV, Ageing Research Laboratories, Campus for Ageing and Vitality, Newcastle University, Newcastle, UK

**Keywords:** Telomeres / Telomere Length, Schizophrenia Spectrum Disorders, Bipolar Disorder, Coffee Consumption

## Abstract

**Objective:** Telomere length is an indicator of cellular ageing, with patients with severe mental disorders tending to have shorter telomeres than the general population. Coffee consumption may reduce oxidative stress, helping prevent biological ageing processes like telomeric shortening. The British National Health Service (NHS) advises limiting caffeine intake to 400 mg/day (4 cups of coffee). However, the role of coffee consumption and telomere length in psychiatric populations remains unclear.

**Methods:** This cross-sectional study included 436 participants (schizophrenia spectrum [n = 259] and affective disorders [n = 177]) from the Norwegian TOP study. Leukocyte telomere length (TL) was measured via blood using quantitative real-time Polymerase Chain Reaction (qPCR). Patients self-reported coffee consumption, quantified as cups per day (no coffee, 1-2, 3-4, 5+).

**Results:** An inverted J-shape was found between TL and coffee intake, peaking at 3-4 cups/day before declining after 4 cups (F = 3.29, p = .02). The largest TL difference was between those drinking the highest recommended dose and non-drinkers (F = 6.13, p = .01). Coffee drinkers within the recommended dose had longer TL, comparable to five years younger biological age, adjusted for confounders.

**Conclusion:** Coffee intake within the recommended dose is linked to longer telomeres in severe mental disorders, comparable to five years younger biological age.

**Key Messages:** *What is already known on this topic?:*

- Patients with severe mental disorders tend to have shorter telomere lengths, an indicator of accelerated cellular ageing.
- Coffee consumption has been noted to possess health benefits, which may help prevent telomere shortening.
- The relationship between coffee consumption and telomere length in psychiatric populations remains unclear.

*What this study adds?:*

- Coffee consumption up to 3-4 cups per day, but not exceeding this amount, was associated with longer telomeres in patients with severe mental disorders.
- Patients consuming up to 4 cups of coffee per day had telomere lengths comparable to a biological age five years younger than non-coffee drinkers.

*How this study might affect research, practice or policy?:*

- Our study suggests the importance of further research investigating the role of coffee consumption in biological ageing.

## INTRODUCTION

Individuals with severe mental disorders (SMD) tend to have a lifespan fifteen years shorter than their unaffected peers[1]. This premature mortality is linked to a higher incidence of somatic diseases - such as cardiovascular diseases and certain forms of cancer[2,3], conditions often associated with advanced chronological age. To that end, researchers have proposed a potentially accelerated rate of ageing being present in patients with SMD, indexed throughout research by variations in telomere length (TL)[4].

Telomeres are structures located on the ends of human chromosomes, made of repeating TTAGGG nucleotides[5], whose role is to guard DNA during replication. Despite telomere attrition being universal, contemporary research has reported shorter TL in both patients with schizophrenia (SZ) [6,7] and bipolar disorder (BD)[8,9], compared to unaffected age-matched peers. Despite reporting shorter telomeres in SMD, researchers still don’t fully understand the aetiology of the cellular differences between these two groups. Due to telomeres being sensitive to environmental factors[10], one avenue of research has been exploring how diet may impact telomere biology[11]. This paper will focus on examining one widely consumed dietary component - coffee.

Coffee has become one of the most widely and frequently consumed beverages across many parts of the world. Epidemiological reviews have indicated that during the 2021/22 period, an estimated 10.56 billion kilograms of coffee were consumed worldwide[12]. A review of coffee consumption using the UK Biobank indicated that of the 468, 629 participants, 77.9% drank coffee, with 19.5% drinking upwards of 3 cups per day[13].

Coffee has been suggested to confer several physiological health benefits, such as improving cognitive acuity[14], lowering the prevalence of neurodegenerative disease[15], reducing risk of obesity, metabolic syndrome or type 2 diabetes, lowering the prevalence of several types of cancer, as well as reduction in the risk of all-cause mortality[16]. However, despite these potentially positive physiological effects, according to several international health authorities, such as the American Food and Drug Administration (FDA) and the British National Health Service (NHS), it is recommended that individuals do not consume more than 400 mg of caffeine per day[17,18], equating to a maximum of approximately 4 cups of coffee. More than 4 cups per day may have negative effects on physical health. As shown in a review paper by Gardiner et al.[19], coffee consumption may reduce sleep time by 45 minutes as well as reduce sleep efficiency, and the consumption of more than 5 cups a day has been linked to an increased risk of panic attacks in vulnerable individuals[20]. Excessive coffee drinking has been noted as a risk factor for several physical health issues, with consumption above the recommended dose causing symptoms such as insomnia, increased urination, gastrointestinal disorders, muscle tremors, irritability, arrhythmia, and flight of ideas[15]. Individuals with hypertension, children, adolescents, and the elderly may be more vulnerable to the side effects of caffeine consumption[21] and are recommended to have lower daily dose than healthy adults.

Contemporary public health research has started to explore the impact coffee consumption may have on other biological processes, such as telomere shortening. Studies of coffee consumption and TL in the general population have provided mixed results, with some indicating a negative association, particularly for instant coffee[22]. However, other studies have suggested a potential protective effect of coffee consumption on TL[23], largely stemming from the proposed antioxidant qualities that coffee may possess[24].

There exists limited research on such behaviours in individuals with SMD, however studies have indicated that individuals with psychiatric disorders ingest significantly higher caffeine levels compared to healthy controls[25]. Further differences have been found between different disorders, with some studies noting that individuals with BD were found to drink more coffee compared to those with SZ, with both psychiatric cohorts ingesting more coffee compared to healthy controls[26]. Moreover, coffee drinking may be exacerbated by smoking, due to the increased metabolism of caffeine stimulated by nicotine[27], with higher rates of smoking being noted in individuals with SMD compared to unaffected controls[28].

However, despite evidence suggesting a faster rate of telomere attrition in SMD, as well as overall higher reported rates of coffee consumption and smoking in psychiatric populations, there is a lack of literature examining such an association. To our knowledge, this is the first study in the literature to investigate the association between coffee consumption and TL in people with schizophrenia or affective disorders. Considering previous research in the general population, we anticipate that moderate coffee consumption will be associated with having longer TL, adjusting for confounders.

## METHODS

### Participants

The study encompassed 436 participants (schizophrenia spectrum [SZ; n = 259] and affective disorders [Bipolar type 1 =114, type II=39, Bipolar not otherwise specified (NOS)=8, and major depressive disorder with psychosis=16, n = 177]), selected from the Norwegian Thematically Organised Psychosis (TOP) study, collected between 2007 and 2018[29]. Participants were recruited from four psychiatric units across Oslo, Norway. Participants were excluded based on the following criteria: Age outside the 18-65 range, not fluent in Norwegian, having a current or past organic psychosis, a history of moderate or severe head trauma, or a somatic disease interfering with brain functioning. All participants provided informed consent. The current study cohort is a subsample of the larger TOP cohort, excluding participants based on available telomere and coffee consumption data (for a further breakdown see Figure S1).

### Clinical Assessment

Patient diagnoses were assessed using the Structured Clinical Interview for DSM-IV (SCID-I, chapters A-E)[30]. The assessments were administered by trained physicians, psychiatrists, and clinical psychologists, all of whom underwent clinical training based on the SCID 101 training program from University College Los Angeles (UCLA)[31]. Overall, the assessment team inter-rater reliability score was between 0.92 and 0.99, proving satisfactory.

Reviews of medical records provided information regarding the Daily Defined Dose (DDD) of psychotropic medication participants were receiving at the time. Information regarding current coffee use was gathered through clinical interviews, by asking participants about how much coffee they currently consume each day (e.g.: How much coffee do you currently drink per day?). Four response choices were available: Zero, 1-2 cups, 3-4 cups, and 5 or more cups. Similarly, clinical interviews aimed to establish smoking habits by asking participants whether they smoked, and if yes, how many years they had been smoking.

### Telomere Length

Telomere length, collected from peripheral blood leukocytes, was assessed in all participants using a quantitative real-time polymerase chain reaction (qPCR)[6,32]. The analysis was carried out on a 384-well plate Applied Biosystems 7900HT Fast Real Time qPCR, beginning with 10 ng of extracted leukocyte DNA being combined with 5 µl of SYBR®Green JumpStart Taq Ready Mix and 0.25 µl of ROX reference dye. The primers used for the telomeric reaction included: 300 nM TelA (5′-CGG TTT GTT TGG GTT TGG GTT TGG GTT TGG GTT TGG GTT-3′) and 900 nM TelB (5′-GGC TTG CCT TAC CCT TAC CCT TAC CCT TAC CCT TAC CCT-3′). The primers used for the singe copy gene (36B4) were: 200 nM 36B4F (5′-CAG CAA GTG GGA AGG TGT AAT CC 3′) and 400 nM 36B4R (5′-CCC ATT CTA TCA ACG GGT ACA A-3′). In addition, three pre-measured DNA samples (10.4 kb, 3.9 kb and 2 kb) were run alongside the study reaction as a control batch. Inconclusive samples as well as outliers (top and bottom 5%) were re-analysed. Analyses of variation coefficients in the study sample noted an intra-assay coefficient of 6.07 % and an inter-assay coefficient of 6.08 %. Overall the qPCR provided researchers with a telomere to single copy ratio (T/S ratio). This was used to estimate mean telomere length, with smaller T/S ratios indicating shorter mean telomere length. All blood samples used in the study were stored in The Biobank - Oslo, Norway.

In addition to the qPCR analysis which yielded the T/S ratio, the authors estimated differences in base pair attrition. This was carried out using the suggested average of 70 base-pair reduction per year as a quantitative estimate of years of accelerated aging[33]. In order to estimate TL attrition, authors subtracted the base pair difference between different groups (e.g.: coffee drinkers - non drinkers) and divided the difference by 70. This provided them with an estimate of years of accelerated ageing.

### Statistical Analysis

All statistical analyses were performed using IBM SPSS software, version 29 (SPSS Inc., Chicago, IL, USA) For categorical variables (sex, ethnicity, coffee consumption), chi-square tests were used to compare distribution between the coffee drinking groups. The remaining comparisons were performed using an ANCOVA. The study only included participants with complete datasets. Any missing data is highlighted in Table 1.

**Table 1.** Demographic Overview.

| Variable | No Coffee<br>(n = 51) | 1-2 cups/day<br>(n = 123) | 3-4 cups/day<br>(n = 118) | 5+ cups/day<br>(n = 144) | Statistics | Post-hoc Test |
| --- | --- | --- | --- | --- | --- | --- |
| Age (years), mean (SD) | 26.35 (9.21) | 27.85 (8.72) | 30.44 (11.01) | 31.81 (9.87) | $F = 5.86$,<br>$p < 0.001$ | 5+ Cups > No<br>Coffe, 1-2 Cups |
| Sex |  |  |  |  |  |  |
| Male, N (%) | 27 (52.9%) | 71 (57.7%) | 61 (51.7%) | 79 (54.8%) | $\chi^2 = 0.95$,<br>$p = 0.81$ | - |
| Ethnicity |  |  |  |  |  |  |
| European, N (%) | 39 (76.5%) | 88 (71.5%) | 97 (82.2%) | 134 (93.1%) | $\chi^2 = 22.19$,<br>$p < 0.001$ | European < Non-European |
| Psychiatric Diagnosis |  |  |  |  |  |  |
| Schizophrenia, N (%) | 22 (8.5) | 74 (28.6) | 69 (26.6) | 94 (36.3) | $\chi^2 = 7.73$,<br>$p = 0.05$ | Schizophrenia <<br>Affective<br>Disorder |
| Affective Disorder, N (%) | 29 (16.4) | 49 (27.7) | 49 (27.7) | 50 (28.2) |  |  |
| Telomere Length (T/S ratio), mean (SE) | 1.06 (0.04) | 1.09 (0.02) | 1.16 (0.02) | 1.15 (0.02) | $F = 3.02$,<br>$p = 0.03$ | - |
| Duration of Smoking, mean (SD) | 4.40 (6.29) | 7.05 (7.59) | 7.68 (8.06) | 13.30 (9.77) | $F = 20.99$,<br>$p < 0.001$ | 5+ Cups > No<br>Coffe, 1-2 Cups,<br>3-4 Cups |
| Lithium Dose (mg/day), mean (SD) | 0.10 (0.32) | 0.10 (0.31) | 0.14 (0.42) | 0.05 (0.26) | $F = 1.19$,<br>$p = 0.31$ | - |
| Antipsychotic Dose (mg/day), mean (SD) | 0.49 (0.65) | 2.02 (6.45) | 1.24 (2.77) | 1.64 (3.41) | $F = 1.59$,<br>$p = 0.19$ | - |
| Antidepressant Dose (mg/day), mean (SD) | 0.29 (0.77) | 0.45 (1.05) | 0.45 (0.85) | 0.43 (0.89) | $F = 0.36$,<br>$p = 0.78$ | - |
| Mood Stabilizers Dose (mg/day), mean (SD) | 0.23 (0.43) | 0.25 (0.47) | 0.27 (0.58) | 0.23 (0.44) | $F = 0.18$,<br>$p = 0.90$ | - |

\*Data on medication was available for 404 participants (92.7% of the sample)

To investigate differences in TL between groups (1 = “no coffee consumption”, n = 44; 2 = “one to two cups per day”, n = 117; 3 = “three to four cups per day”, n = 110; or 4 = “five cups or more per day”, n = 133), we performed ANCOVAs adjusting for age, sex, ethnicity, years of tobacco use and medication use (Daily Defined Dose [DDD] of lithium, antidepressants, mood stabilisers, and antipsychotics) with Bonferroni post hoc corrections. Due to a skewed distribution, TL values were log-transformed. Sensitivity analyses were performed, adjusting for diagnostic group status as well as comparing non-coffee drinkers and those drinking 1-4 cups per day, adjusted for confounders (see above).

Lastly, in order to assess whether the relationship between coffee consumption and telomere length differed by sex or diagnosis, an interaction analysis was conducted using ANCOVA. Coffee consumption and sex were included as fixed factors, while age, ethnicity, medication DDD, years of smoking, and diagnosis were included as covariates. The same process was repeated for the diagnostic group, adjusting for age, sex, ethnicity, medication DDD, and years of smoking. The models tested the main effects and the Coffee × Sex and Coffee x Diagnosis interactions.

## RESULTS

### Demographic overview

Table 1 provides the descriptive statistics for the study sample (n = 436), distributed between the four coffee drinking groups. In summary, the group drinking 5+ cups was significantly older than the groups drinking no coffee and 1-2 cups per day (F = 5.86, p < 0.001). While there was no difference between the groups in terms of sex, all four groups were comprised of significantly more Europeans than non-Europeans (χ^2^ = 22.19, p < 0.001). Moreover, individuals with schizophrenia imbibed significantly more coffee than individuals with an affective disorder (χ^2^ = 7.73, p = 0.05). There were no significant differences between the four groups in terms of psychotropic medication use. In total, 77.3% of the sample smoked (n = 337), with the average duration of smoking use for the whole sample being nine years (M = 8.98, SD = 8.93). Further post-hoc analyses indicated that the 5+ cups group smoked for significantly longer than the other four groups (F = 20.99, p < 0.001)

### Coffee intake and telomere length

ANCOVA results indicate a significant difference in TL between different coffee consumption groups (*F* = 3.29, *p* = 0.02), with the distribution forming an inverted J-shape (see Figure 1). Bonferroni adjusted post-hoc tests revealed significant differences in TL between the group of no coffee consumption (n = 44) and the group consuming 3-4 cups, (n = 110; *p* = 0.04). Analyses were adjusted age, sex, ethnicity, number of years of using tobacco, and medication Daily Defined Dose (DDD). The association remained when sensitivity analyses were performed adjusting for diagnostic group status (*F* = 3.21, *p* = 0.02).

**Figure 1.**
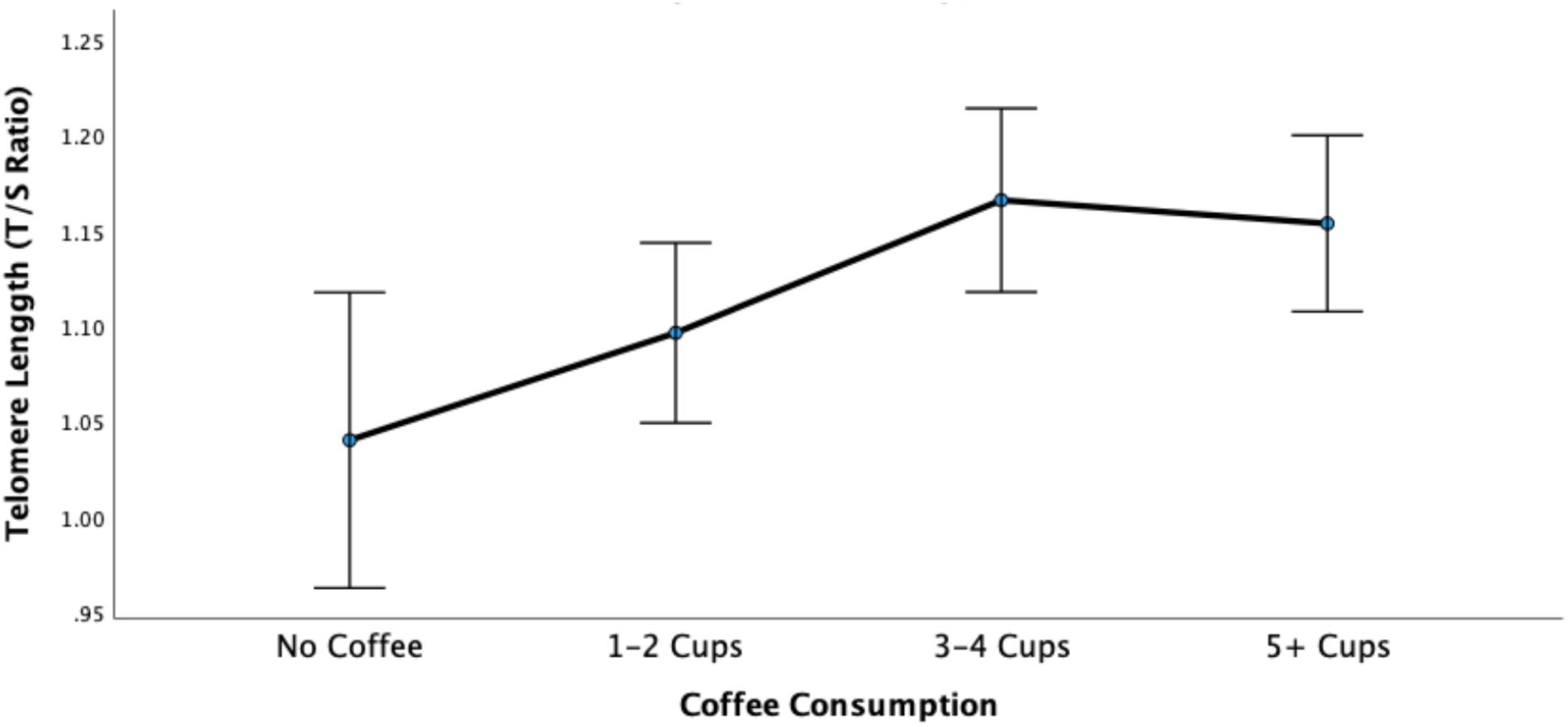
Coffee consumption and telomere length ANCOVA, *F* = 3.29, *p* = .02. Analysis adjusted for age, sex, ethnicity, years of tobacco use, and medication (Daily Defined Dose [DDD] of lithium, antipsychotics, antidepressants, and mood stabilisers). The error Barrs indicate 95% confidence intervals.

Individuals who reported no coffee consumption (n = 44) had shorter TL than patients consuming coffee within the recommended number of cups per day (n = 227; *F* = 6.13, *p* = 0.01). Based on an average of 70 base-pair reductions per year[33], this represents five years younger biological age in the coffee-drinking group, adjusted for confounders (for confounders, see above). Both interaction analyses for sex (*F* = 0.84, *p* = 0.47) and diagnosis (*F* = 0.62, *p* = 0.60) were not statistically significant, indicating that the effect of coffee consumption on telomere length did not significantly differ between males and females, or individuals with schizophrenia and affective disorders.

## DISCUSSION

Our findings indicate an inverted J-shape association between coffee consumption levels and cellular ageing as measured by TL in patients with a severe mental disorder, adjusted for age, sex, ethnicity, medication, tobacco use, and group status. Specifically, coffee consumption up to the recommended limit of four cups per day was associated with longer telomere lengths that could represent up to five years younger biological age compared to non-coffee drinkers, adjusted for relevant confounders.

The positive association between coffee intake and TL observed in our study is in line with previous large scale studies conducted in individuals without a mental disorder. Specifically, both the British Nurses’ Health Study[23]as well as the American National Health and Nutrition Examination Survey (NHANES)[34], indicated longer telomeres in individuals who drank coffee, compared to those who abstained. However, as noted previously, adverse effects of coffee have also been noted, namely in a large scale UK Biobank study conducted by Wei et al.[22], wherein coffee consumption was associated with decreases in TL. Moreover, the NHANES study further highlighted that while coffee intake overall was associated with longer TL, increased intake of caffeine specifically was associated with reductions in TL[34]. Similar trends were noticed in our own study, particularly with individuals who exceeded the daily recommended intake of 4 cups/day, showing a decrease in TL.

There are multiple mechanistic pathways which could be underpinning the association highlighted in our study. Firstly, coffee has been noted to contain several bioactive compounds such as chlorogenic acid (CGA), cafestol, kahweol, trigonelline, and melanoidins[35], all of which possess potent antioxidant properties. Particularly, CGA as well as trigonelline have been noted to play an important role in promoting the activity of the Nrf2 pathway, a crucial antioxidant pathway within the body[24,36]. Interestingly, studies of patients with schizophrenia have noted a downregulation of this specific pathway[37,38]. Moreover, CGA and other bioactive compounds found in coffee have also been noted to reduce the formation of pro-inflammatory cytokines, helping reduce inflammation[39]. Chronic and systemic inflammation are a common pathophysiological feature found in SMD[40,41], highlighting the potential protective effects that coffee could have in this population. In addition, telomeres are highly sensitive to both oxidative stress and inflammation[42], further highlighting how coffee intake could help preserve cellular ageing in a population whose pathophysiology may be predisposing them to an accelerated rate of ageing. However, despite the potential benefits of coffee, as noted in our study findings, consuming more than the daily recommended amount of coffee may also cause cellular damage and TL shortening through the formation of reactive oxygen species[43].

Another key mechanistic pathway worth considering is the Akt/GSK3β/β-catenin pathway and the influence it has on the expression level of a key telomerase enzyme, TERT. The Akt/GSK3β/β-catenin pathway is a regulatory pathway associated with cellular proliferation, survival, and differentiation[44]. It’s cascade includes Akt (Protein Kinase B) phosphorylating and thereby deactivating GSK3β (Glycogen Synthase Kinase 3 Beta), which allows for the accumulation of β-catenin[44]. β-catenin is highly important for regulating gene expression, including *TERT*, a key subunit of the telomerase complex, that can extend telomere length and thus is crucial for telomere maintenance[45]. In-vitro studies of cancer cells have found that caffeine treatment downregulated GSK3β activity through phosphorylation[46]. Similarly in-vivo animal studies have noted that chronic caffeine treatment upregulated phosphorylated Akt, and downregulated GSK3β in mice[47]. In addition to this particular pathway, in-vitro studies have also noted that caffeine treatment has been found to increase *TERT* gene promoter activity, which consequently lead to increases in *TERT* mRNA, and longer TL in caffeine treated cells[48]. These findings are highly relevant in the context of our previous research, wherein we noted both reduced *TERT* expression as well as shorter TL in individuals with SZ, compared to healthy controls[32]. In effect, these findings suggest the potential cellular benefits that coffee consumption may have in promoting healthy ageing in individuals with SMD. In addition, we have to take into consideration other previously highlighted behaviours found in SMD, which may be influencing coffee consumption patterns. Most notably, individuals with SMD have been noted to possess higher rates of smoking compared to unaffected controls[28]. This is particularly relevant, as tobacco-associated chemicals, such as nicotine, may up-regulate the production of liver enzymes (e.g.: CYP1A2) associated with caffeine metabolism[27]. Studies in the normative population have indicated higher serum levels of this enzyme[49], as well as a faster metabolism of caffeine[50] in smokers compared to non-smokers. The significantly higher coffee consumption in smokers compared to non-smokers was also observed in our sample, suggesting that smoking may influence coffee drinking in SMD as well.

### Study limitations

One of the main limitations of our study was the robust measurement of coffee consumption. The current self-report data inquired only about the number of cups of coffee ingested per day, and not time of the day consumed or instant versus filter coffee, which have been shown to influence the link between coffee consumption and health[16,22]. We also did not have information on other sources of caffeine (e.g.: tea, energy drinks, soda) or the caffeine concentration of cups of coffee drank, which could have provided different results.

Moreover, whilst it is our speculation that TL was increased due to potential conferred antioxidant/ anti-inflammatory properties, we did not have data on peripheral antioxidant/inflammation levels. In addition, our study only included information on psychotropic medication use, and not alternative medication that participants may have been using for somatic illnesses (e.g.: beta blockers, statins, metformin, etc.).

In addition, the current study is only comprised of a psychiatric sample (affective disorder & schizophrenia) without a healthy control comparison group, and is conducted cross-sectionally, limiting the ability to specify the directionality of our hypothesis.

Furthermore, TL was measured using qPCR giving a mean TL measure and not proportion or number of short telomeres within a sample[6]. Although this is a validated measure of TL often used in the literature, we cannot rule out that measuring number of critically short TL may have given additional information about the role of coffee relative to markers of biological ageing. We also included only one marker of biological ageing (TL) while ideally several markers should have been included (including epigenetic clock, brain age etc).

Our study is to our knowledge the first study to show an inverted J-shaped association between TL and coffee consumption in patients with a severe mental disorder, suggesting that, in moderation, coffee consumption might have a positive effect but has a reverse effect in large doses. Coffee consumption up to the limit of recommended cups per day was associated with a five-year’s younger biological age compared to non-coffee drinkers, adjusted for confounders. This was not statistically significant in individuals consuming coffee above the recommended dose. Our study contributes to a new understanding of both the potential protective and detrimental effects of coffee consumption on TL in a psychiatric population, deserving further attention. As people with severe mental disorders tend to have high coffee consumption, our study suggests potential health benefits by monitoring coffee consumption to reduce intake above the recommended daily dose.

## Data Availability

All data relevant to the study are included in the article or uploaded as supplementary information.

## CONTRIBUTORSHIP

VM and MA wrote the first draft of the manuscript. All other authors - MDF, EH, DS, IA, SD, CMR, DQ, VB, NES, MO & OA - contributed to the study. VM is the guarantor of this study.

## ACKNOWLEDGEMENTS

We thank the patients who took part in the study and the NORMENT researchers who contributed to the data collection.

## ETHICAL APPROVAL

This study was approved by the Regional Committee for Medical Research Ethics and the Norwegian Data Inspectorate (Code: 2009/2485/REK sør-øst).

