## Supplementary figures and images for "Coffee intake is associated with telomere length in severe mental disorders"

### Supplementary Figure 1

**Figure S1.** Flowchart of participants exclusion

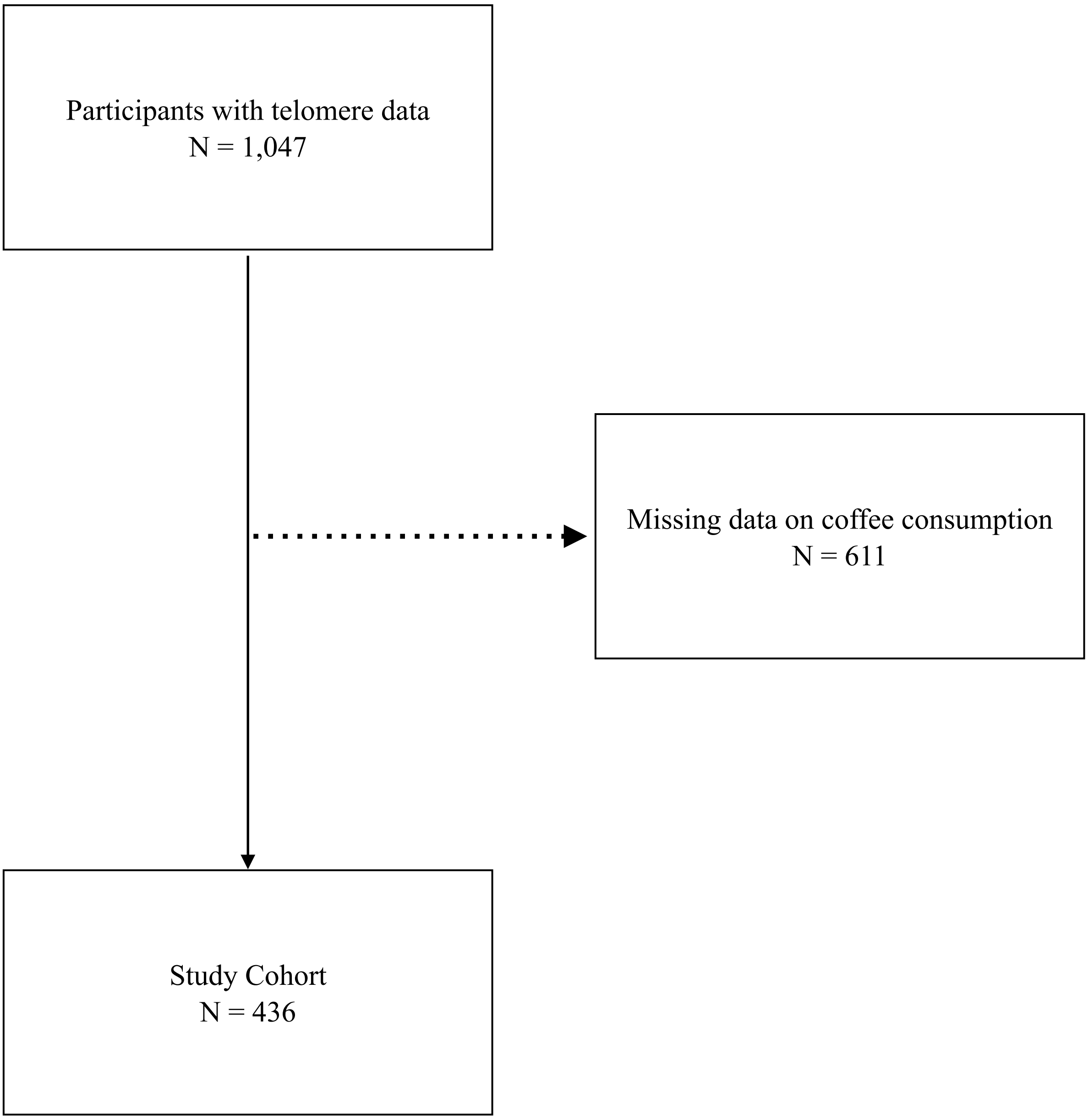
